# Network-based analysis of genome-wide biobank data boosts discovery of genetic associations in psoriasis

**DOI:** 10.64898/2025.12.05.25340137

**Authors:** Giann Karlo Aguirre-Samboní, Gwenaëlle Lemoine, Julio Molineros, Florian Massip, Chloé-Agathe Azencott

**Affiliations:** CBIO-Centre for Computational Biology, Mines Paris, PSL University, 60 boulevard Saint-Michel, Paris, F-75006, France; Institut Curie, PSL Research University, 26 rue d’Ulm, Paris, F-75005, France; INSERM, U1331, 26 rue d’Ulm, Paris, F-75005, France; Johnson and Johnson Innovative Medicine, 1400 McKean Rd, Spring House, Pennsylvania, 19002, United States of America

**Keywords:** Network-based methods, psoriasis, biological networks, GWAS

## Abstract

Psoriasis is a common autoimmune disease with a strong genetic component. Targeting the *IL-23/IL-17* pathways for the treatment of moderate-severe psoriasis has proven successful, which makes it a good benchmark for other drug discovery approaches. Genome Wide Association Studies (GWAS) identify genetic loci associated with a phenotype (e.g. disease). However, using identified loci to understand the disease mechanisms remains challenging. In response, gene networks methods that consider gene-gene relationships have been developed. In a previous study, we showed that the combination of network methods improved the power, reproducibility, and interpretability of those approaches on a small cohort of breast cancer patients. The goal of the present study is to benchmark the performance of the same combination of network methods on a psoriasis cohort from the UK biobank (with a particular interest on the *IL-23/IL-17* pathways). By applying this method, we identified well known psoriasis genes, demonstrating the viability of our approach. Our method further identified a small set of genes of the MRPL family robustly associated with psoriasis, demonstrating that this approach can provide insights into the potential disease mechanisms. Our study also shows that our method is applicable to large-scale biobank data to produce novel findings, even in well-studied diseases with strong genetic signal.

## 1 Introduction

Genome-wide association studies (GWAS) analyze thousands of genotypes to identify single nucleotide polymorphisms (SNPs) that are associated with complex traits across different phenotypes. GWAS output groups of correlated SNPs that each demonstrate a statistically significant association with the trait being studied. Results from GWAS can be applied in various ways, including enhancing our understanding of the biology underlying specific phenotypes, making clinical risk predictions (Uffelmann et al. 2021), and identifying potential therapeutic targets (Buniello et al. 2019). While GWAS have successfully uncovered thousands of genetic variants associated with common diseases, they face challenges due to the high dimensionality of genomic data. First, the number of variants vastly exceeds the number of samples, which limits statistical power and restricts detection to variants with relatively large effect sizes. Second, most phenotypes are highly polygenic, influenced by thousands of SNPs with small, additive effects, making it difficult to distinguish true signals from background noise (Yang et al. 2010). Third, GWAS typically explain only a fraction of trait heritability, partly due to rare variants, epistatic interactions, and gene-environment effects not captured by standard models (Manolio et al. 2009; Zuk et al. 2012). Thus, despite the promise of large-scale initiatives such as biobanks, fully harnessing GWAS data requires analytical strategies capable of addressing these statistical and biological complexities.

One approach is to integrate GWAS results with biological networks, particularly gene interaction networks. These networks represent genes as nodes and biological relationships — such as co-expression, protein-protein interaction, or shared pathways — as edges. This interconnected structure effectively models the underlying biological mechanisms arising from gene interactions. From this perspective, complex traits and diseases emerge not from individual genes acting alone but from the interrelated action of gene networks (Silverman et al. 2020; Zitnik et al. 2024).

Biological network models help address GWAS limitations in several ways. First, they provide a framework for aggregating weak signals across genes that participate in shared functions, thereby improving power to detect relevant loci. Second, they incorporate prior biological knowledge that can guide interpretation of GWAS hits. Third, they support the local hypothesis—that genes contributing to the same phenotype tend to cluster in the network and form disease modules (Barabási et al. 2011; Furlong 2013). Fourth, they align with both the infinitesimal model of polygenicity (Barton et al. 2017) and the omnigenic model, which posits that core trait-associated genes are influenced by a broader network of peripheral genes through regulatory interactions (Boyle et al. 2017).

Many methods have been proposed over the years to apply network biology principles to GWAS data. Starting from a gene interaction network in which genes have been assigned scores according to their relevance to a phenotype of interest, their goal is to identify modules (*i.e.* group of connected genes) with a large proportion of high scoring nodes (Leiserson et al. 2015; Dittrich et al. 2008; Liu et al. 2017; Wang et al. 2015; Gwinner et al. 2017). Network-based GWAS approaches compute the score of a gene from the p-values of association with the phenotype of SNPs that are mapped to this gene, typically through their proximity to the gene on the DNA sequence (Jia and Zhao 2014; Visonà et al. 2024). These methods rely on “guilt-by-association” (Farber 2013; Lee et al. 2011) to “rescue” genes with a non-significant p-value through their connection with other significant genes (Chimusa et al. 2019; Azencott 2016). However, since these methods formalize the question differently, they yield varying solutions, which raises concerns regarding the reproducibility and reliability of the results obtained. To address this issue, Climente-Gonźalez et al. (2021) proposed a strategy to combine these methods into a consensus network and successfully applied it to a familial breast cancer GWAS dataset. While a classical GWAS had very little power on this dataset, yielding only one or two significant genes, they showed that this consensus network approach significantly improved the power to discover genes associated with a disease. This suggests that the approach is a good way to compensate for lack of power.

Here, we study whether such an approach can be useful in the context of a much better powered GWAS dataset. To this end, we applied this approach to a GWAS on psoriasis—a disease with a strong polygenic basis—from the UK Biobank (UKB) (Bycroft et al. 2018). Psoriasis is a chronic disease characterized by excessive proliferation of keratinocytes and infiltration of immune cells in the dermis and epidermis (Greb et al. 2016). Our dataset integrates genotype data from about 500 000 patients stored in the UKB, along with clinical information. We show that the consensus network obtained by the combination of five different tools, namely HotNet2 (Leiserson et al. 2015), SigMod (Liu et al. 2017), Heinz (Dittrich et al. 2008), dmGWAS (Jia et al. 2011; Wang et al. 2015) and LEAN (Gwinner et al. 2017) with a protein-protein interaction network obtained from BioGRID (Stark et al. 2006; Oughtred et al. 2021), recovers both the psoriasis-related genes typically identified through GWAS, such as *IL-17*, *IL-23* and *TNF* pathways, and genes that have been previously related to psoriasis but not through GWAS, such as *MRPL* genes involved in mitochondrial translation. Ultimately, our research demonstrates the utility of network-based approaches into genetic research, suggesting that they may unlock new avenues for understanding and potentially treating complex diseases such as psoriasis.

## 2 Material and methods

### 2.1 UK Biobank cohort

The UKB is a large-scale, population-based study that collected detailed information from approximately half a million participants in the United Kingdom. The data collected includes biological samples as well as comprehensive health-related and demographic measures (Bycroft et al. 2018).

### 2.2 Phenotype definition

Our study used samples from 193 608 patients that were estimated to have white British ancestry. We defined 10 105 psoriasis cases using the hospital episode statistic ICD-10 codes (L40.x codes) and primary care data codes that included the terms “psoriasis” or “psoriatic” (see full list in table S11). The remaining 183 503 samples were used as controls.

#### 2.2.1 UKB SNP microarray dataset

Genotypic data was assayed on the UKB Axiom Array, an Affymetrix array designed to represent British and European genetic diversity (Bycroft et al. 2018; Affymetrix 2020). The array assayed 784 256 single nucleotide coding variants (SNVs). Our quality control filtered out SNPs with:

‒ Minor Allele Frequency (MAF) lower than 1%,
‒ Minor Allele Count (MAC) lower than 100,
‒ Hardy-Weinberg equilibrium (p-value *<* 1e-15), and
‒ genotype missingness on more than 10% of the samples.

We excluded patients who requested that their data be withdrawn. We reduced spurious associations due to potential population stratification by focusing only on white British ancestry. The final dataset contained 593 068 SNPs.

### 2.3 SNP-level and gene-level analysis

To investigate the association between genotype and psoriasis, we conducted a per-SNP allelic test with 1 degree of freedom using the *χ*^2^ statistic, as implemented in PLINK version 1.9 (Chang et al. 2015).

Then, we used MAGMA (de Leeuw et al. 2015) to derive gene-level association scores from the p-values of the SNPs mapped to their corresponding genes. We decided to use MAGMA over VEGAS, as done in (Climente-Gonźalez et al. 2021) mainly by two considerations: 1) VEGAS exhibited notable computational inefficiency, rendering it impractical for large-scale analyses; 2) VEGAS imposes a truncation threshold on p-values at 1 × 10*^−^*^6^. This means that any p-values smaller than this threshold are arbitrarily set to 1 ×10*^−^*^6^ (Nakka et al. 2016). This p-value truncation presents a critical impediment for network-based analytical approaches, which frequently require the discrimination of highly significant associations with p-values well below the 1 × 10*^−^*^6^ threshold. The inability to accurately quantify these extreme p-values compromises the statistical power necessary for downstream network analyses, potentially leading to the loss of biologically relevant signals and reduced sensitivity in identifying gene-gene interactions or pathway-level associations.

We mapped SNPs to genes (referred to as the annotation step in the MAGMA documentation CTGLAB (2015) version 1.10) using GRCh37 genomic coordinates as reference. Specifically, we mapped SNPs to a gene if they were located within a 50 kb interval of the gene’s boundaries. Subsequently, we computed gene-level p-values, known as the gene analysis step in CTGLAB (2015), by aggregating SNPs at the gene level and testing the joint association of all SNPs within the gene with the phenotype. MAGMA’s gene analysis employs a multiple regression approach to account for linkage disequilibrium among SNPs and to detect multi-marker effects (de Leeuw et al. 2015).

### 2.4 Network methods

The network methods described hereafter use undirected simple graphs to connect nodes and display subnetworks. Then, a *graph* (or *network*) is a tuple *G* = ⟨*V, E* ⟩, where *V* is a finite set of *vertices* (or *nodes*), and 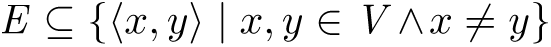 is the *relation* among vertices whose elements are called *edges*. A *subgraph* (or *subnetwork* or *module*) of *G* is a tuple *S_G_* = ⟨*V_S_, E_S_* ⟩, where *V_S_* ⊆ *V* and *E_S_* ⊆ *E* (where we drop the reference to *G* if no confusion can arise).

#### 2.4.1 Protein-Protein Interaction Network

Protein-protein interactions (PPIs) describe crucial relations for the realization of molecular processes in the human body, serving as a key source of therapeutic interventions against diseases (Gao et al. 2023). PPIs can be mathematically represented as *graphs* (or networks), where proteins are nodes and interactions are edges connecting them. These interactions depict specific physical contacts between proteins within cells, which occur at defined binding regions and have distinct biological meanings, thereby serving specific functions (De-Las-Rivas and Fontanillo 2010). Additionally, proteins may be linked if they collaborate towards a common objective in a metabolic or signaling pathway, regulate one another through intermediaries, or collectively contribute to a shared cellular structure (Szklarczyk et al. 2022).

We assume that a gene interacts with another if its corresponding encoded protein exhibits an interaction within the PPI Network (PPIN). We utilized a PPI dataset from BioGRID (Stark et al. 2006; Oughtred et al. 2021; the BioGRID Team 2025) as input. We filtered the PPIN by taxonomy ID 9606 to include only interactions involving *Homo sapiens*. We then leveraged the filtered PPIN to apply the various subnetwork-search algorithms to display potential gene modules related to psoriasis.

#### 2.4.2 Subnetwork-search algorithms

Genes that contribute to the same biological function in a PPIN tend to be topographically proximate and can be related in different manners, such as densely interconnected modules, nodes surrounding a hub, through a linear path, and so forth (Yook et al. 2004; Gursoy et al. 2008). When designing a network-based method, several factors must be considered, including the scoring of nodes, number of connections per node, constitution of many connected components or singletons, and computational efficiency. Numerous approaches have been proposed in literature for subnetworks detection within biological networks. In this work, we use five subnetwork-detection algorithms, all of them exploiting the PPIN. We selected open-source methods for which implementation and documentation were available. For these methods, we employed the default parameter values or those recommended by their respective authors.

Network-based methods for identifying disease-associated gene modules each employ unique strategies to balance connectivity, association strength, and computational efficiency: SigMod (Liu et al. 2017) and Heinz (Dittrich et al. 2008) target globally optimal or densely connected subnetworks—SigMod via a graph mincut approach that tunes the trade-off between signal strength and interconnectivity (using parameter *λ*), and Heinz through integer-linear programming to solve a Prize-Collecting Steiner Tree (PCST) problem, maximizing node scores while preserving connectivity. dmGWAS (Wang et al. 2015; Jia et al. 2011) and LEAN are more heuristic and GWAS-focused, with dmGWAS employing a seed-and-extend approach to iteratively build modules from GWAS-significant seeds—often producing overlapping regions—while LEAN (Gwinner et al. 2017) detects “star-shaped” subnetworks by testing for p-value enrichment among a gene’s immediate neighbors, resulting in a conceptually simple and statistically conservative approach. HotNet2 (Leiserson et al. 2015), distinctively, uses a diffusion-based algorithm to distinguish “hot” (high-scoring) from “cold” genes, propagating node scores (e.g., p-values) through the network via an insulated heat diffusion process to uncover both known and novel subnetworks, particularly in cancer. While SigMod and Heinz prioritize dense or high-scoring modules, dmGWAS and LEAN provide scalable, GWAS-tailored alternatives, and HotNet2 bridges local and global perspectives by leveraging diffusion to reveal less obvious but biologically relevant subnetworks, often missed by purely connectivity-driven or seed-based methods.

#### 2.4.3 Consensus network

We compiled all the different gene networks yielded by the methods into a *consensus network*. The consensus network addresses the inherent heterogeneity of all methods by computing their overlap. Different assumptions are taken for conceiving how a subnetwork is identified; thus we filtered genes so they would only remain in the consensus network if they were selected by at least three of the five different methods (Fig. 1). The stringency of this filter is higher than that of Climente-Gonźalez et al. (2021) where they keep only genes selected by two out of six methods.

**Fig. 1:**
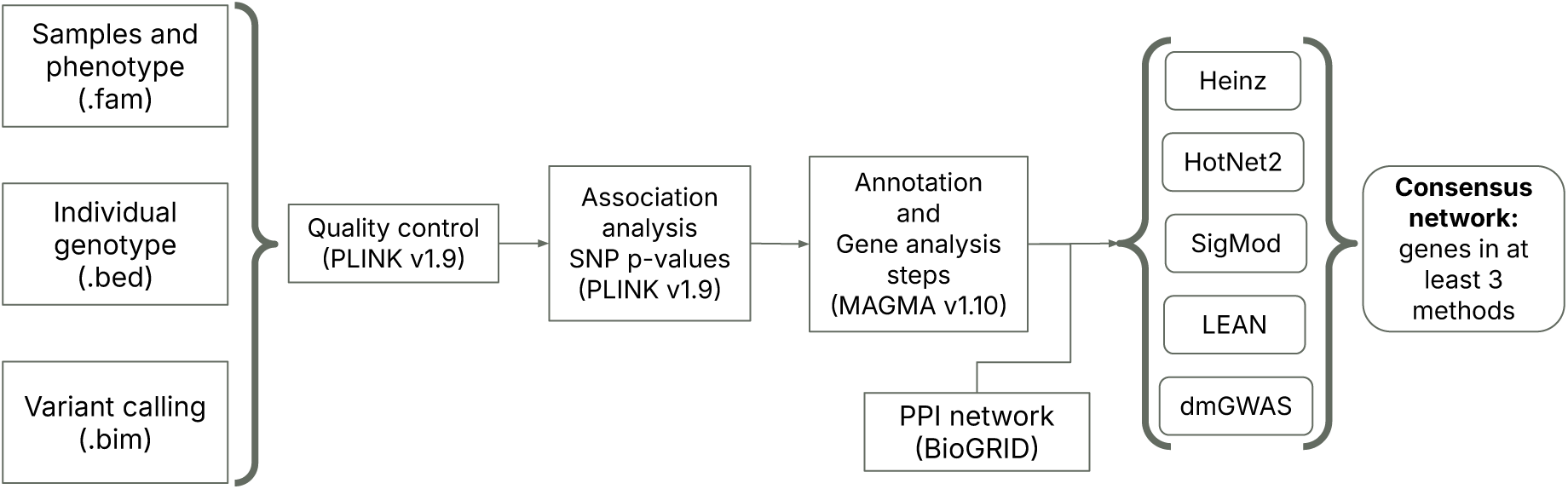
General workflow for constructing the consensus network.

**Fig. 2:**
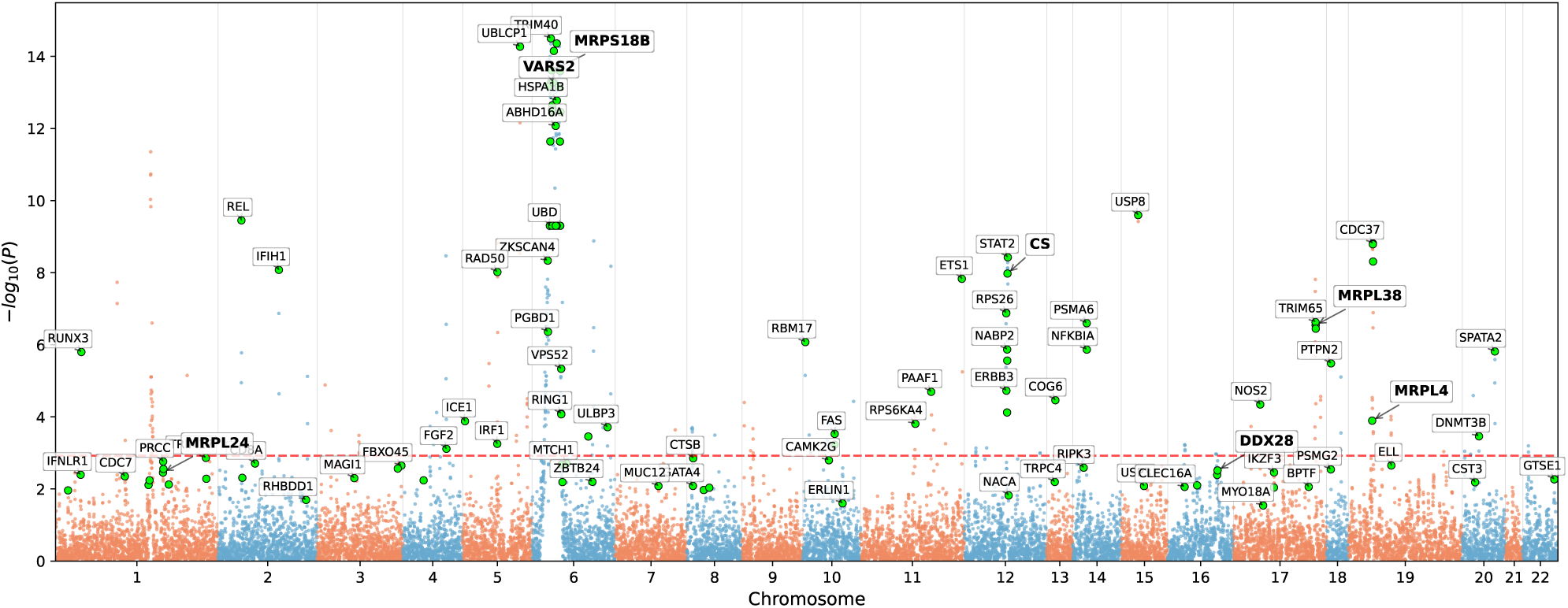
Manhattan plot of genes; in color and with labels some of the genes in the consensus network (128 genes or green data points in total). The red line indicates the Benjamini-Hochberg threshold (1.19 × 10*^−^*^3^ for a significance threshold of 0.05). The names of some genes connected in the consensus network are bold, although not all are significant and are dispersed throughout the genome.

**Fig. 3:**
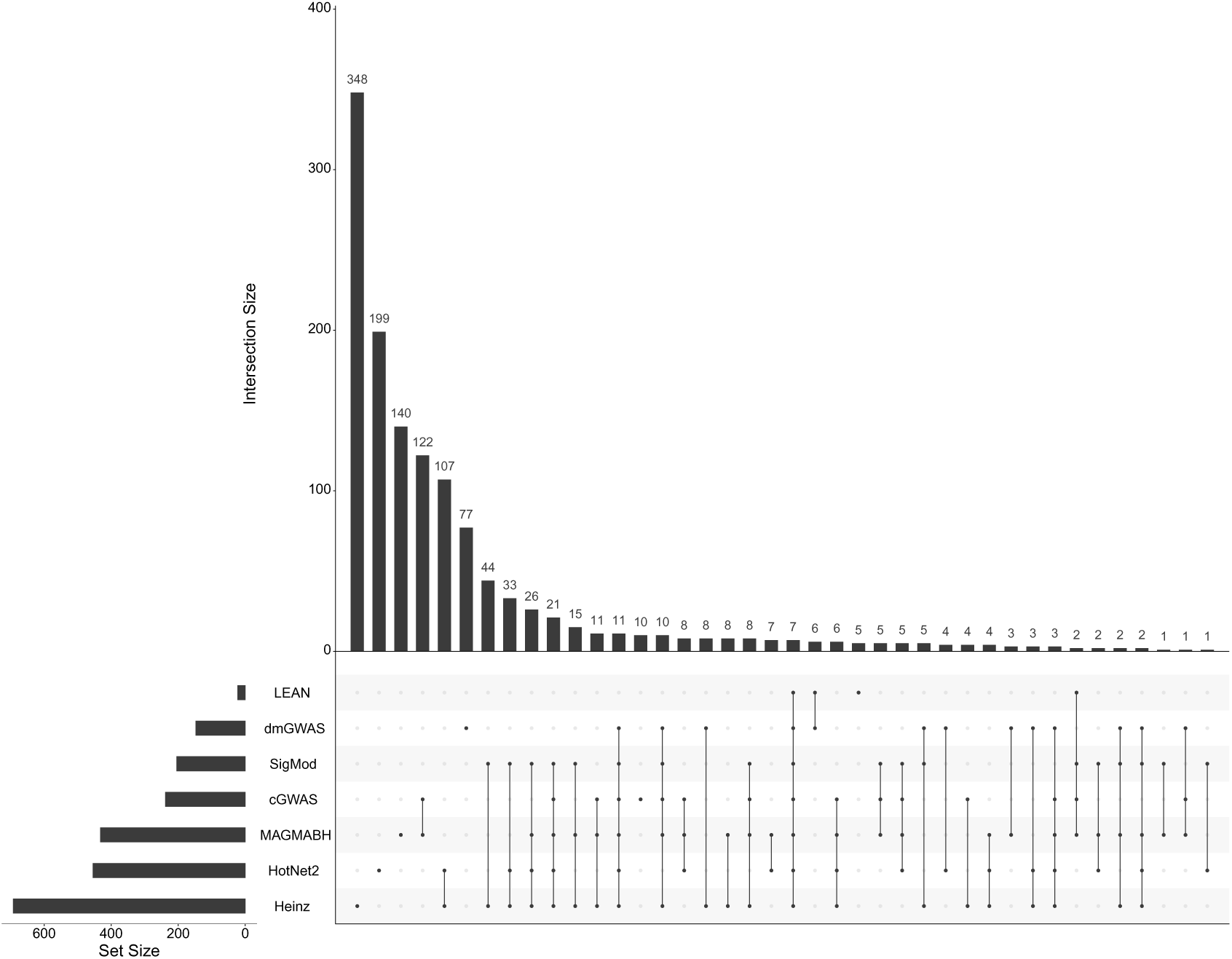
UpSet plot for representing all the common genes across different intersections of gene sets produced by the network-based methods. MAGAMBH corresponds to gene p-values outputted by MAGMA after applying the Benjamini-Hochberg procedure, cGWAS corresponds to gene p-values mapped from SNP p-values after the association test computed using PLINK v1.9.

#### 2.4.4 Stable consensus network

Following Climente-Gonźalez et al. (2021), we built a *stable consensus network* to address the inherent instability of the network methods. Indeed, iterations of the pipeline on different subsets of the input data yield different gene lists. So as to establish a more robust network, we aggregate the genes most frequently selected across 25 distinct solutions, generated by applying the five network-based methods to five different subsets (5-fold subsampling setting) of 80% of the samples (see Fig. S3 for an illustration of this pipeline). This stable consensus network aims to identify subnetworks that are frequently altered, thereby enhancing its resilience to noise and instability. Additionally, the stable consensus approach still accounts for the heterogeneity of each method by evaluating their results across various datasets. We followed the criterion established in Climente-Gonźalez et al. (2021), where the choice of keeping genes selected at least 7 times out of 30 resulted in 68 genes. In our case, we have a total of 25 solutions, and keeping genes selected at least 7 times retained 258 genes, which is approximately 13% of all genes selected at least once (Fig. S4).

### 2.5 Pathway enrichment analysis

We searched for potentially enriched pathways across modules and the consensus network. To do this, we included modules of interest in separate queries. The background gene set was chosen as the intersection of the gene analysis step outcome (see Section 2.3) and the PPIN (the BioGRID Team 2025). We used the Enrichr online tool because of: 1) its extensive coverage, comprising approximately 450 000 annotated gene sets organized into about 220 gene set libraries, and 2) its support for programmatic access, which enables rapid execution of analyses, visualization of results in multiple formats, and storage of queries for subsequent reference (Chen et al. 2013; Kuleshov et al. 2016; Xie et al. 2021). From the report given by the online tool, we downloaded the table for pathways enriched in the Reactome database (Milacic et al. 2023) and kept only significantly enriched pathways with two or more genes in overlap (*>* 1), and adjusted p-value *<* 0.05.

### 2.6 Code and data availability

We implemented a computational pipeline to facilitate the execution of steps in the GWAS analyses, including the gene annotation of SNPs, computation of gene scores, and the application of the five distinct network methods. Our pipeline is designed to be versatile and can be applied to any GWAS dataset, enabling the efficient execution of these processes, https://github.com/giannkas/gwas-bionets2/. We have no authorization nor agreements to share genotype data publicly for confidentiality and privacy reasons.

## 3 Results

### 3.1 SNP-level and gene-level analyses

We identified 2 708 significantly psoriasis-associated SNPs (*P <* 5×10*^−^*^8^) using PLINK 1.9 for 593 068 SNPs after quality control (see Methods, Section 2.2.1). 2 584 significant SNPs (95.42% of 2 708) mapped to at least one gene, and we annotated 240 genes (1.24% of 19 427 genes in GRCh37/hg19 (Church et al. 2011; Consortium 2009)), with at least one significant SNP mapped to each of them. These annotated genes are labeled as *cGWAS* in Fig. 4.

**Fig. 4:**
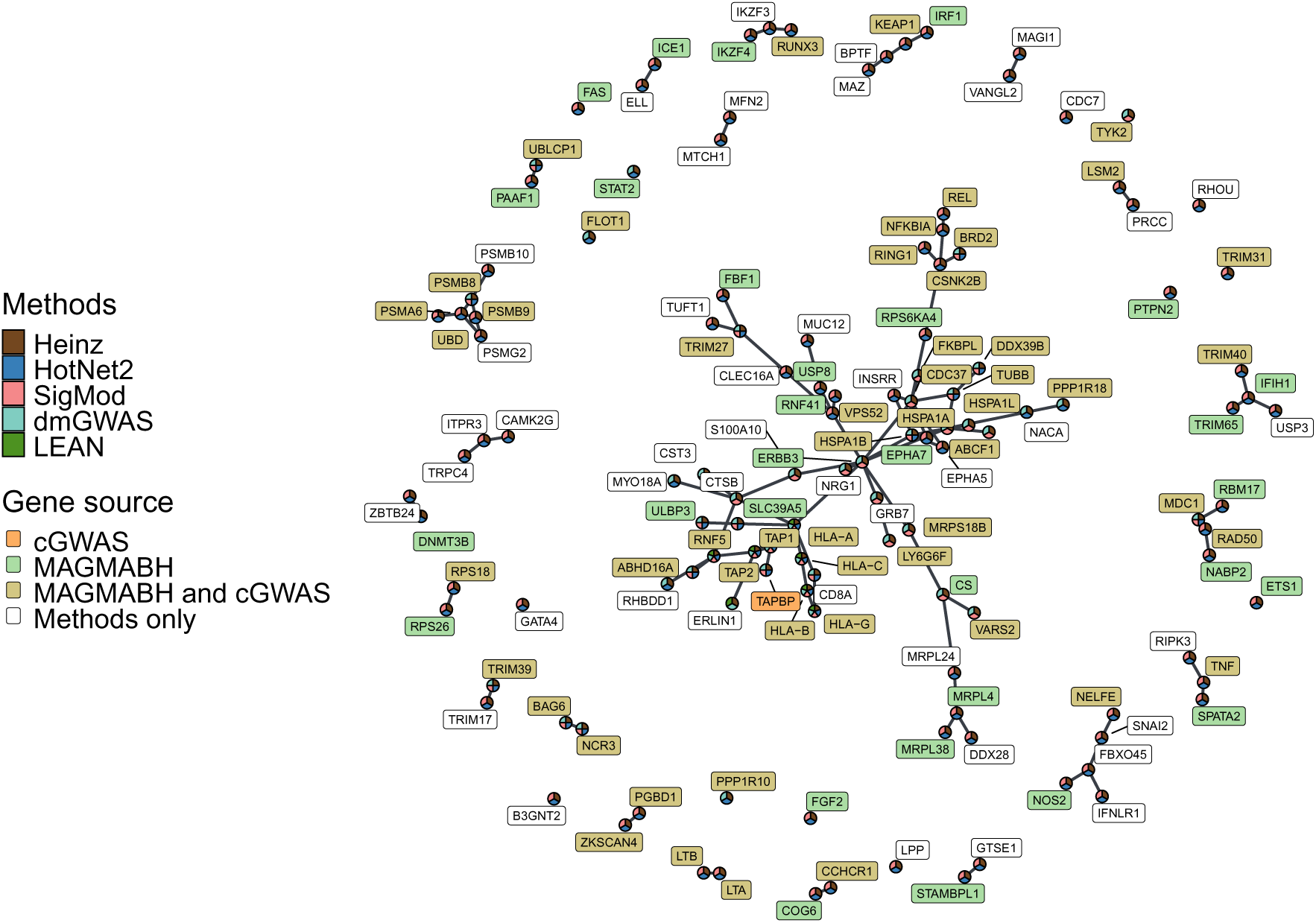
Consensus network built by choosing those genes that were selected in at least 3 of the 5 methods. The network contains 128 nodes corresponding to genes. Methods and genes are highlighted to depict which method (Methods, pie chart) selected one particular gene, and if a gene (Gene source, colored labels) was also found significant in classical GWAS (cGWAS), or by MAGMA analysis after Benjamini-Hochberg correction (MAGMABH).

Moreover, we used all 593 068 SNP p-values after the allelic association tests to annotate and compute gene p-values. We annotated 18 399 genes (94.70% of 19 427 genes in GRCh37/hg19 (Church et al. 2011; Consortium 2009)), meaning that at least one SNP was mapped to each of them. A substantial portion of SNPs in the array (401 659, 67.73% of 593 068) mapped to at least one gene.

Gene-wise analysis with MAGMA yielded 18 086 genes, of which 434 passed the gene-wise significance thresholds of *P <* 0.05 and *FDR <* 1.19 × 10*^−^*^3^. We found 11 837 genes in the BioGRID PPI network. Among these, 10 327 (or 87.24%) of the human genes are in common between the PPI network and the MAGMA output. MAGMA p-values were used as the initial scores for the network methods (Fig. 1). Fig. 2 shows a Manhattan plot of the p-values of these genes. Genes that passed the gene-wise significance thresholds are labeled as *MAGMABH* in Fig. 4.

### 3.2 Subnetwork identification

The algorithms rely on gene p-values to identify interesting subnetworks (or subgraphs, or modules) of the PPI network. The number of genes selected by each method varied from 22 to 693 (Heinz: 693, HotNet2: 454, SigMod: 204, dmGWAS: 147, LEAN: 22). Fig. 3 compares all the genes selected by the five methods, with genes that passed the Benjamini-Hochberg correction (MAGMABH), and the annotated genes from significant SNPs through a classical GWAS approach (cGWAS). Note that because network approaches exclude isolated significant genes (272 genes, including only MAGMABH or cGWAS fields), none of the different methods selected one of them as part of the consensus. Additionally, network-methods allowed for the selection of non-significant genes (840 genes, excluding MAGMABH or cGWAS fields) that were missed by GWAS and MAGMA approaches.

While Heinz (Dittrich et al. 2008) sought the highest-scored subnetwork and LEAN (Gwinner et al. 2017) searched “star-shaped” subnetworks with one central gene, HotNet2 (Leiserson et al. 2015), SigMod (Liu et al. 2017), and dmGWAS (Wang et al. 2015) identified strongly connected subnetworks or modules within the PPI network likely associated with psoriasis (Fig. S1 shows the topology produced by the five different network-based methods). In contrast, most significant genes in cGWAS and MAGMABH appeared alone, which complicates functional interpretation and justifies the use of network methods.

### 3.3 Consensus network

We built a consensus among the five methods by selecting only those genes identified by at least three methods (Fig. 4). The resulting consensus network contains 128 genes, 44 (34.4% of all selected genes) of which were exclusively selected by network-based methods. These genes allowed us to identify modules of psoriasis-related genes that would not have been detected otherwise. The consensus network contains three modules, the largest one containing 43.7% of all selected genes, including *TNF*, *NFκB* and *HLA* genes. The *NFκB-HLA* module connects highly interconnected genes (such as *ERBB3* associated genes) with multiple cGWAS or MAGMABH genes. This module appears as such because network methods have incorporated non-significant genes that serve to connect significant genes. For example, the genes *CTSB, CLEC16A, S100A10, RIPK3, INSRR, ERLIN1*, and *GRB7* —which connect to major histocompatibility complex (MHC) genes such as *TAP2*, *HSPA1A*, and *HLA*, as well as *TNF* —were selected exclusively by the network methods.

Additionally, our results highlight the role of a module of mitochondrial ribosomal proteins (*MRPL* and *MRPS*, Fig. 4). Although some of these genes were already identified as significant by MAGMABH (*MRPS18B, CS, VARS2, MRPL38* and *MRPL4*, see Fig. 2), MRPL-family genes identified in the consensus network are connected together because *MRPL24* was selected by three network-based methods (Heinz, HotNet2 and SigMod, see Fig. 4). Nevertheless, there are 61 genes that do not belong to modules with more than four connected nodes (Fig. 4), suggesting they are consensus genes in distinct possible modules estimated by different network methods.

We also evaluated how well the consensus network recovered known psoriasis genes (Fig. 5). Genes associated with psoriasis in previous meta-GWAS (Tsoi et al. 2017; Ran et al. 2019; Dand et al. 2025; Zhang et al. 2025), as well as pathways involving *IL-17*, *IL-23*, *TNF*, *IL-1*, *IFN-γ*, *IFN*-mediated, *IL-22*, *IL-6*, *IL-9*, *NF-κB*, *JAK-STAT*, *MAPK*, *PI3K-AKT* and *WNT* genes (Tsoi et al. 2012; Belinky et al. 2015; Girolomoni et al. 2019; Nair et al. 2009; Guo et al. 2023) were present in the consensus network. The consensus network included genes in the *IL-23* /*IL-17* axis (central to psoriasis pathogenesis). These pathways drive Th17 cell activation, neutrophil recruitment, and keratinocyte hyperproliferation (Dhillon-LaBrooy et al. 2024; Liu et al. 2020). Tumor Necrosis Factor (*TNF*) amplifies inflammation by inducing pro-inflammatory mediators (e.g., growth factors, adhesion molecules, and chemokines), which promote immune cell infiltration into the skin and synergize with *IL-17* /*IL-23* signaling (Scarpa et al. 2006; Baliwag et al. 2015). Together, these cytokines create a feed-forward loop that sustains chronic inflammation and epidermal hyperplasia, contributing to the cytokine-rich milieu characteristic of psoriatic lesions (Liu et al. 2020; Menter et al. 2021).

**Fig. 5:**
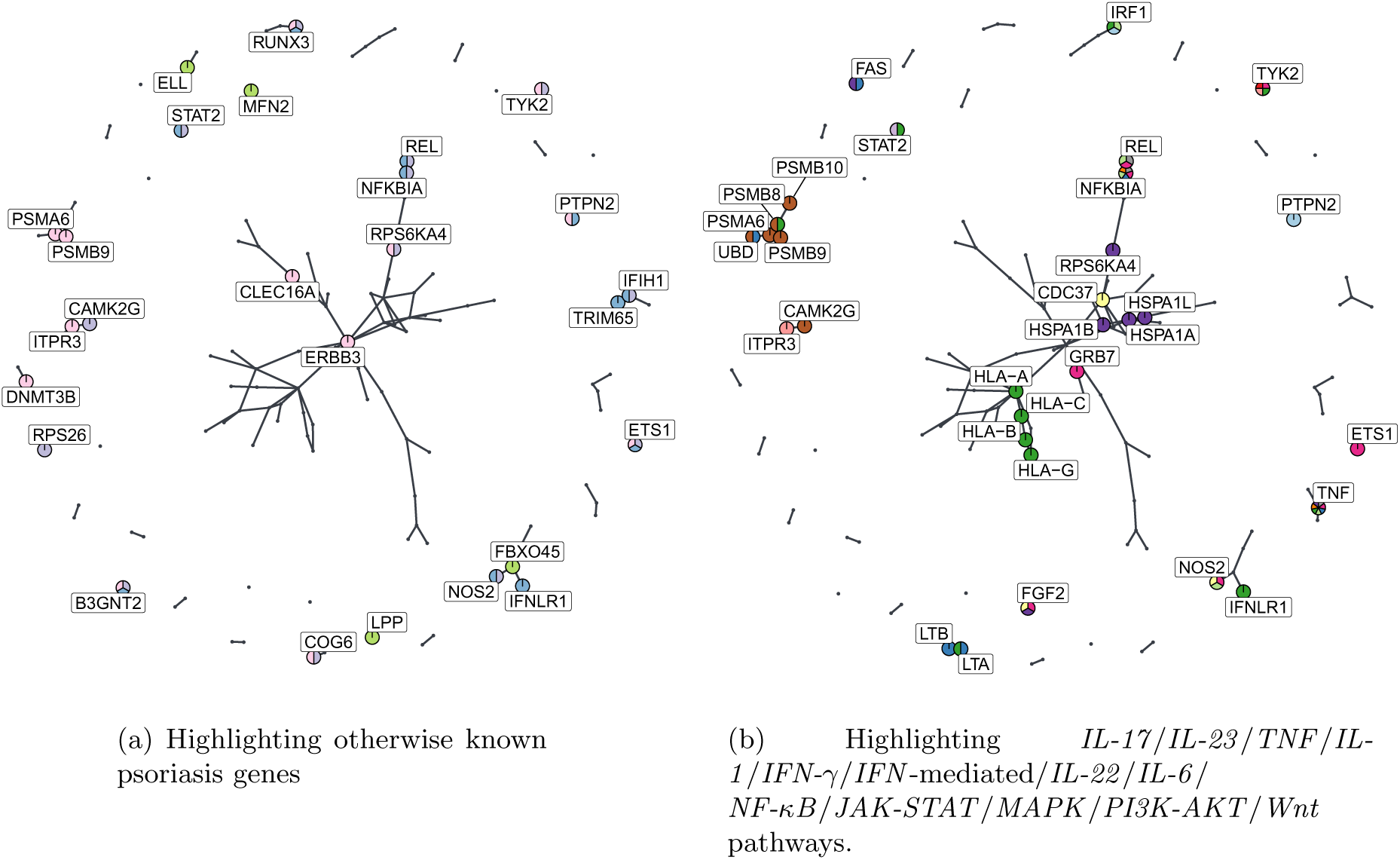
Consensus network comprising 128 genes built by choosing those genes selected in at least 3 methods out of 5. The first network (a) highlights genes in common with known gene lists from Ran et al. (2019)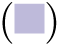, Tsoi et al. (2017) 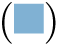, Dand et al. (2025) 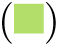 and Zhang et al. (2025) 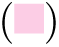. The second network (b) highlights genes in common with *IL-17* 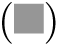, *IL-23* 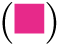, *TNF* 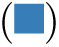, *IL-1* 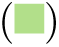, *IFN-γ* 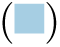, *IFN*-mediated 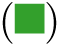, *IL-22* 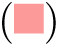, *IL-6* 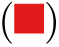, *NF-κB* 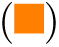, *JAK-STAT* 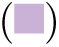, *MAPK* 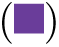, *PI3K-AKT* 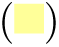 and *WNT* 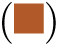 pathways.

### 3.4 Stable consensus network

We evaluated the robustness of each method to data perturbation. To this end, we built a stable consensus network using a similar approach to Climente-Gonźalez et al. (2021). We kept genes selected in at least 7 out of 25 solutions as shown in the resulting stable consensus network (Fig. S4) made of 258 genes. Of the total of 128 genes selected in the consensus network, 119 genes (92.96%) are also present in the stable consensus network, indicating that the majority of these genes are stable. Fig. S2 shows the stable consensus network, overlaid with sets of genes with strong evidence for involvement in the pathogenesis of psoriasis.

Many of these psoriatic genes were identified in the stable consensus network (81 genes), and in the consensus network (44 genes). We found 43 psoriatic genes in common between the consensus network and the stable consensus network: *MFN2, HLA-G, IFIH1, ERBB3, FAS, CDC37, HSPA1A, NOS2, NFKBIA, TNF, PSMB8, PSMA6, ETS1, REL, RPS26, FGF2, DNMT3B, STAT2, RPS6KA4, IRF1, ELL, HLA-A, HLA-C, CAMK2G, ITPR3, LTA, UBD, LTB, RUNX3, PTPN2, FBXO45, TYK2, PSMB9, PSMB10, HLA-B, HSPA1B, COG6, HSPA1L, LPP, TRIM65, B3GNT2, IFNLR1, CLEC16A* (Tsoi et al. 2012; Dand et al. 2025; Girolomoni et al. 2019; Nair et al. 2009; Ran et al. 2019; Zhang et al. 2025). The *GRB7* gene was only present in the consensus network and is part of *IL-23* pathway. The remaining 38 genes in the stable consensus network were also within the same pathways and meta-GWAS-psoriatic genes as in the consensus network. *IL-9* pathway did not appear in either of the two networks.

We found no additional relevant modules in the stable consensus network beyond those already identified in the consensus network. Furthermore, as previously mentioned, approximately 92.3% of the genes in the consensus network are stable. The consensus network effectively detected associated subnetworks within the larger module of the stable consensus network. Therefore, unless otherwise specified, we will focus on the consensus network for the remainder of the paper.

### 3.5 Gene set enrichment analysis

We performed gene set enrichment analyses using Reactome Pathways (Milacic et al. 2023) for each of the 3 modules of the consensus network (containing at least five genes), as well as for the whole consensus network. As expected, the whole consensus network shows strong enrichment in pathways involving genes in the 6p21 and 6p22 regions such as *HLA-A, HLA-B, HLA-C, HLA-G, HSPA1A, HSPA1B, HSPA1L, PSMB8, PSMB9, TAP1, TAP2, TAPBP, NCR3* and *TRIM31* confirming expected associations between these pathways and psoriasis etiology (Greb et al. 2016) (see Tab. S6 and Tab. S5).

One of the small modules, which contains five genes (*NELFE, SNAI2, FBXO45, IFNLR1*, and *NOS2*), shows no enrichment that meets our criteria (refer to Methods, Section 2.5). The second module, made of six genes, namely *PSMB10, PSMB9, PSMB8, PSMA6, UBD* and *PSMG2*, is enriched for the following pathways: Proteasome Assembly, Post-translational Protein Modification and Metabolism of Proteins (see Tab. S2 and Tab. S1 for further details).

The largest module in the consensus network contains 56 genes, and is enriched for several pathways in common (22 in total) with the full consensus network (see Tab. S4 and Tab. S3). A total of 16 significant pathways were found exclusively within the 56-gene module, while 14 significant pathways were identified solely in the complete consensus network. Among the 16 pathways found only in the 56-gene module, four do not contain genes from the MHC: ERBB2 Activates PTK6 Signaling, ERBB2 Regulates Cell Motility, PI3K Events in ERBB2 Signaling and SHC1 Events in ERBB2 Signaling—all identified by the same two genes: *ERBB3* and *NRG1*. The remaining 12 pathways are composed either of a majority of genes located in the 6p21 or 6p22 cytogenic bands (HSF1 Activation, Class I MHC Mediated Antigen Processing & Presentation, Neutrophil Degranulation, HSP90 Chaperone Cycle for Steroid Hormone Receptors (SHR) in the Presence of Ligand, Mitochondrial Unfolded Protein Response (UPRmt), ABC-family Proteins Mediated Transport, PKR-mediated Signaling, RHOBTB2 GTPase Cycle), or of interactions involving a greater number of genes outside these regions. For instance, the genes *MRPL4* (19p13.2), *MRPL24* (1q23.1), *MRPL38* (17q25.1), and *MRPS18B* (6p21.33) (Safran et al. 2021) had strong enrichment in pathways related to mitochondrial function (Table S3 and Table S4), more specifically, in mitochondrial translation. To our knowledge, the association between these pathways and psoriasis has never been proposed. Nevertheless, previous studies have identified a relationship between mitochondrial function, mitoribosome genes, and psoriasis (Therianou et al. 2019; Alwehaidah et al. 2022; Dhillon-LaBrooy et al. 2024; Huo et al. 2025; Gu et al. 2015). These studies employed a variety of methods, including gene expression analysis, mitochondrial protein quantification, measurement of circulating mitochondrial DNA—as a biomarker for psoriasis—, functional inhibition of mitochondrial translation, and Mendelian-randomization studies.

Furthermore, we also performed gene set enrichment analysis for the biggest module in the stable consensus network and the full network itself (see Tab. S7, Tab. S8, Tab. S9, Tab. S10). In particular, both analyses yielded the same list of pathways enriched in either the 215-gene module or the full network, indicating that the isolated genes incorporated by the full stable consensus network offer no additional biological interpretation. We conducted exhaustive comparisons between the 56-gene module, the consensus network, and the stable consensus network to assess shared and non-shared pathways, including those involving genes in the MHC region as well as genes outside this region (see Tab. S12 - Tab. S19). The comparison between the consensus network and the stable consensus network identified 23 pathways in common, while 13 and 33 pathways were exclusively detected in each network, respectively. Although the 56-gene module did not share as many pathways with the stable consensus network as it did with the consensus network, it still demonstrated overlap, with 14 pathways shared between the two, while 24 and 42 pathways were enriched individually in each network, respectively.

## 4 Discussion

The ability of GWAS to uncover the underlying mechanisms of complex diseases has been subject to discussion (Boyle et al. 2017). A key challenge relates to the omnigenic model, which proposes that gene functions are interconnected within dense regulatory networks. According to this model, as GWAS sample sizes grow, widespread pleiotropy will be detected, which may offer limited mechanistic insight. Conversely, the traditional statistical framework of GWAS can be overly conservative and underpowered, hindering the discovery of novel associations.

To address these limitations, several network-based methods have been developed. They leverage both association scores and pre-established interaction networks to contextualize the biological relationships between genes and SNPs. The network methods we employ are genecentric and can be applied to various types of biological networks, reflecting the inherent biological functions that arise from biomolecular interactions. In this study, we applied five of such methods to SNP-array data from the UK Biobank, focusing on psoriasis as the phenotype. We evaluated the effectiveness of a consensus network approach in analyzing a large dataset with substantial genetic signals. In other words, we determined whether combining different network-methods for identifying gene modules in a protein interaction network can enhance the identification of genes associated with psoriasis, compared to classical GWAS.

All five methods—Heinz, HotNet2, SigMod, dmGWAS, and LEAN—produced compelling results (Fig. S1), allowing for the identification of gene modules or subnetworks. In some cases, the consensus of these methods revealed genes associated with biological pathways that had no prior connection to psoriasis. Notably (Fig. 4), seven genes—*DDX28, MRPL38, MRPL4, MRPL24, CS, VARS2*, and *MRPS18B* —were selected by three methods. Five of these seven genes remained significant after Benjamini–Hochberg correction of MAGMA gene p-values. *DDX28* and *MRPL24*, however, were identified only by the network methods, which enabled their connections to the other five genes; this underscores the added value of network-based approaches for genotype–phenotype association in well-powered datasets with strong genetic signals. As detailed in Section 3.5, *MRPL4, MRPS18B, MRPL38* and *MRPL24* are enriched in mitochondrial-related pathways, including Mitochondrial Translation Initiation, Mitochondrial Translation Elongation, Mitochondrial Translation Termination, and overall Mitochondrial Translation, based on Reactome annotations (Milacic et al. 2023).

While prior psoriasis research has implicated individual ribosomal proteins (e.g. *RPL9, RPL3, RPS8, RPL11, RPL22, RPS3A, RPL7A, RPS5, RPS27, EEF1A1*) (Liu et al. 2024; Swindell et al. 2015; Zeng et al. 2021) and ribosome biogenesis (Liu et al. 2025) in psoriasis progression, the coordinated association of mitochondrial ribosomal proteins *MRPL4, MRPS18B, MRPL38* and *MRPL24* as a functional set related to the mitochondrial translation machinery appears to reflect a novel finding.

These observations are not entirely unprecedented: UK Biobank PheWeb analyses (Gagliano et al. 2020) suggest that *MRPL4* and *MRPS18B* lie in a region nominally associated with psoriasis. Additionally, mitochondrial dysfunction (Huo et al. 2025), mitochondrial translation signaling in *V γ*4 + *T* cells (Dhillon-LaBrooy et al. 2024), and mtDNA secretion (Therianou et al. 2019) have been proposed as potential contributors to psoriasis risk. Nevertheless, these four mitochondrial ribosomal genes have not previously been collectively associated with psoriasis pathogenesis.

DEAD-Box Helicase 28 (*DDX28*) has been shown to be downregulated in psoriatic arthritis compared with healthy controls (Stoeckman et al. 2006). *DDX28* belongs to a family implicated in immune signaling (Rao and Mahmoudi 2022) and plays an essential role in early stages of mitoribosome large-subunit biogenesis (Tu and Barrientos 2015). *VARS2* has been associated with psoriasis in a Mendelian-randomization study of individuals of European ancestry (Li et al. 2025), and in a GWAS of a Taiwanese population (Yang et al. 2024). *VARS2* encodes mitochondrial valyl-tRNA synthetase, a key enzyme in mitochondrial protein synthesis that catalyzes the attachment of valine to its corresponding transfer RNA during mitochondrial translation (Uhĺen et al. 2015).

To our knowledge, citrate synthase (*CS*) has not been directly associated with psoriasis. We propose that any potential association could arise from altered citrate and tricarboxylic acid (TCA) cycle metabolite pools in immune-mediated inflammatory diseases—for example, reduced urinary citrate levels in patients with higher disease activity in psoriatic arthritis (Alonso et al. 2016; Koussiouris et al. 2021). Perturbation of the citrate–acetyl-CoA axis (via ATP-citrate lyase, *ACLY*) might connect mitochondrial citrate to lipid synthesis (Rodriguez et al. 2012) and histone acetylation (Lozoya et al. 2019), processes modulated by keratinocyte hyperproliferation and *IL-17* –driven inflammation (Kao et al. 2025; Kulkoviene et al. 2025).

We identified several meta-GWAS studies that reported genes associated with psoriasis as well as signaling pathways implicated in psoriasis pathogenesis. Collectively, these gene lists and pathway annotations comprised a total of 18 independent sources used to validate our findings. In the consensus network, 44 of 128 genes overlapped with one or more of these validated sources (Fig. 5). Some of these 44 genes formed smaller subnetworks separated from the large central module; several of those contained at least five connected nodes and warranted enrichment analysis. For example, we performed gene-set enrichment analysis on the *NELFE-SNAI2-FBXO45-NOS2-IFNLR1*, but the result did not meet our criteria for significance. Nonetheless, *FBXO45* appears in Dand et al. (2025), *IFNLR1* is implicated in interferon-mediated pathways relevant to psoriasis (Tsoi et al. 2017; Guo et al. 2023), and *NOS2* is reported in two studies (Tsoi et al. 2017; Ran et al. 2019), and participates in *IL-23*, *IL-1* and *PI3K-AKT* signaling.

*NELFE* (Negative Elongation Factor Complex Member E) is a subunit of the NELF complex located in the MHC region at 6p21.33 (Safran et al. 2021). The complex is essential for macrophage activation and inflammatory responses; NELF deficiency has been shown to reduce *IL-10* production and attenuate proinflammatory cytokine expression (Yu et al. 2020), suggesting a plausible link to psoriasis. *SNAI2* (Snail Family Transcriptional Repressor 2, also known as *SLUG*) plays a critical role in keratinocyte function relevant to skin inflammatory disorders, regulating epidermal progenitor cell states and differentiation (Mistry et al. 2014). *SNAI2* is induced via the *TLR3-SLUG-VDR* axis after skin injury, enhancing wound healing by modulating *IL-36γ* expression—an axis relevant to psoriasis pathogenesis (Tortola et al. 2012; Jiang et al. 2017; Guo et al. 2023). Although primarily studied in wound healing, *SNAI2* influences inflammatory responses through regulation of *IL-36* family members, which are elevated in psoriatic skin (Jiang et al. 2017).

Another small module comprising previously identified psoriatic genes includes *TRIM40, IFIH1, TRIM65*, and *USP3*. Meta-GWAS studies support associations for *TRIM65* (Tsoi et al. 2017) and *IFIH1* (Ran et al. 2019; Tsoi et al. 2017). *TRIM40* is linked with the risk of guttate psoriasis (Wang et al. 2025) and functions as an inhibitor of *NFκB* activation (Yang and Xia 2021); the *TRIM40* variant *rs757262* exhibits varying risk profiles across autoimmune diseases (Cagliani et al. 2011). *USP3* (Ubiquitin Specific Peptidase 3) is notable for its deubiquitinase and histone-binding activities (Bethesda MD). Deubiquitinases modulate inflammatory responses (Li et al. 2021); *USP3* reduces reduces type I interferon (*IFN-I*) responses (Cui et al. 2013), and the *IL-36/IFN-I* axis has been implicated in extracutaneous inflammation in psoriasis (Catapano et al. 2020).

In this study, we used data derived from SNP array genotyping, which limits our analysis to variants captured by this technology. The UK Biobank additionally provides whole-exome sequencing data (Backman et al. 2021; Szustakowski et al. 2021) and whole-genome sequencing data (Carss et al. 2025), encompassing a large number of common and rare variants. A promising direction is to extend the proposed pipeline to incorporate rare variants, with the aim of identifying more biologically informative pathways and assessing the robustness of the workflow when applied to larger and more heterogeneous variant sets. Numerous methods for incorporating rare variants into GWAS have been proposed (e.g., Lee et al. (2014); Goswami et al. (2021)). Incorporating rare variants may further enhance subnetwork identification within biological networks and reveal additional biologically relevant pathways.

Network-based methods formalize the “guilt-by-association” assumption in different ways, producing distinct but credible outcomes. Thus, our findings may be influenced by the particular methods chosen. Many additional network-based GWAS methods have been proposed recently (Zhu et al. 2021; Muzio et al. 2023; Yilmaz et al. 2021; Carlin et al. 2019; Reyna et al. 2018; Visonà et al. 2024), and integrating them into our pipeline could further enhance analysis.

The choice of PPIN can also significantly impact the final output, since different databases compile different interactions. We used BioGRID (Stark et al. 2006; Oughtred et al. 2021; the BioGRID Team 2025) database. Using a different PPIN source could yield different insights. For instance, Huang et al. (2018) found that larger networks were often more useful for identifying disease-related genes, whereas smaller but more specific networks—such as tissue-specific interactions from the Integrated Interactions Database (Kotlyar et al. 2015)—might be preferable in other settings. Other network types, for example tissue-specific coexpression networks (Sabik et al. 2021), could also be used. A comprehensive evaluation of multiple interactomes is a worthwhile subject for future work.

Overall, this study confirms the usefulness of network-based GWAS in providing a more comprehensive understanding of the genetic basis of complex traits than classical GWAS, even in relatively well-powered datasets.

## Data Availability

All data produced in the present work are contained in the manuscript.

