## Supplementary figures for "Network-based analysis of genome-wide biobank data boosts discovery of genetic associations in psoriasis"

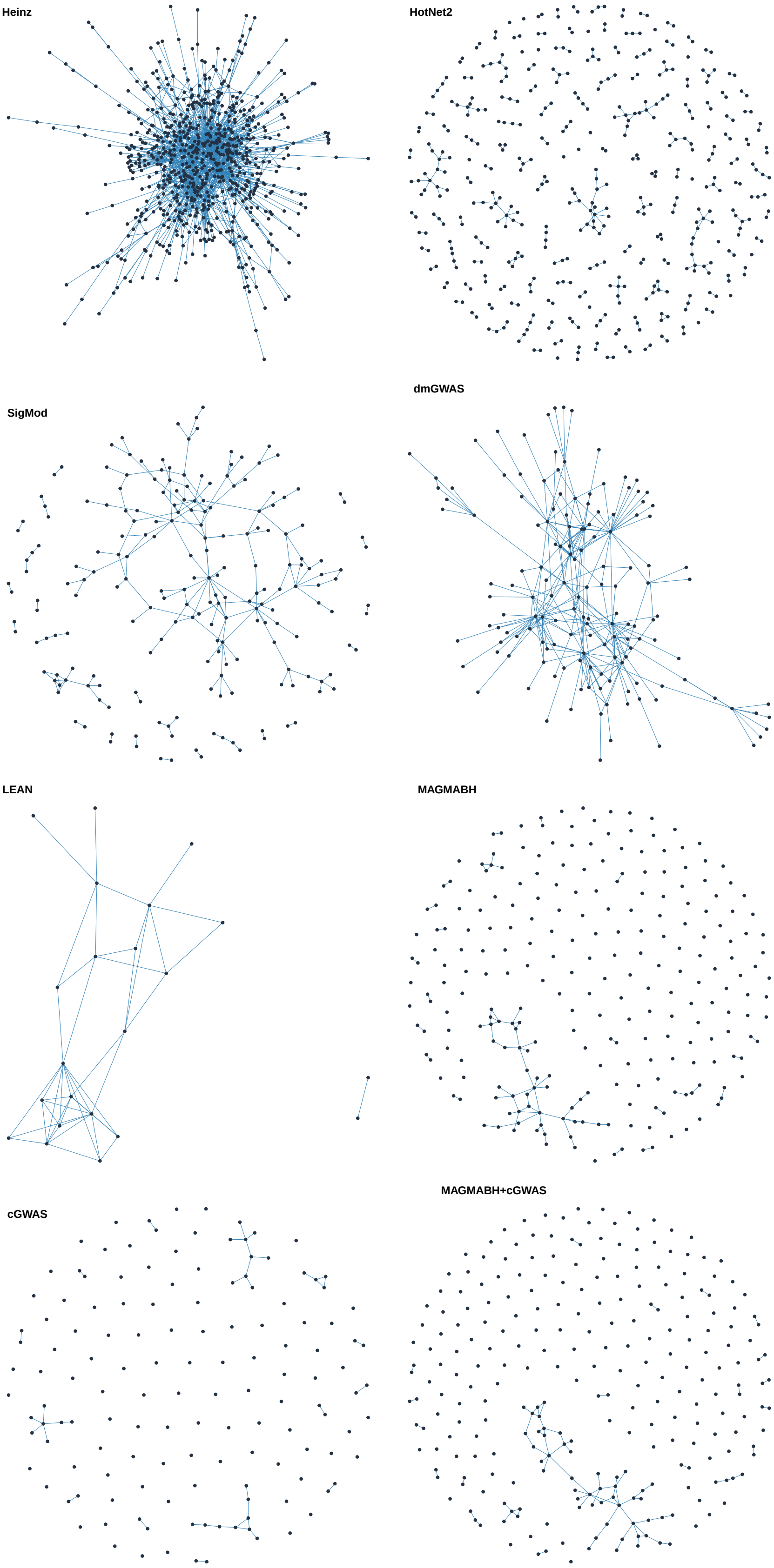


**Fig. S1**: Networks or undirected simple graphs produced by **Heinz** (Dittrich et al. 2008), **HotNet2** (Leiserson et al. 2015), **SigMod** (Liu et al. 2017), **dmGWAS** (Wang et al. 2015), **LEAN** (Gwinner et al. 2017), **MAGMABH** which corresponds to MAGMA output (de Leeuw et al. 2015) after Benjamini-Hochberg correction, **cGWAS** (classical GWAS) which corresponds to mapping significant SNPs to the corresponding gene, **MAGMABH+cGWAS** which corresponds to joining both genes from MAGMABH and cGWAS. Heinz selected 693 genes; HotNet2 selected 454 genes; SigMod selected 204 genes; dmGWAS selected 147 genes; LEAN selected 22 genes; MAGMABH selected 432 genes; cGWAS selected 240 genes; MAGMABH+cGWAS selected 448 genes.

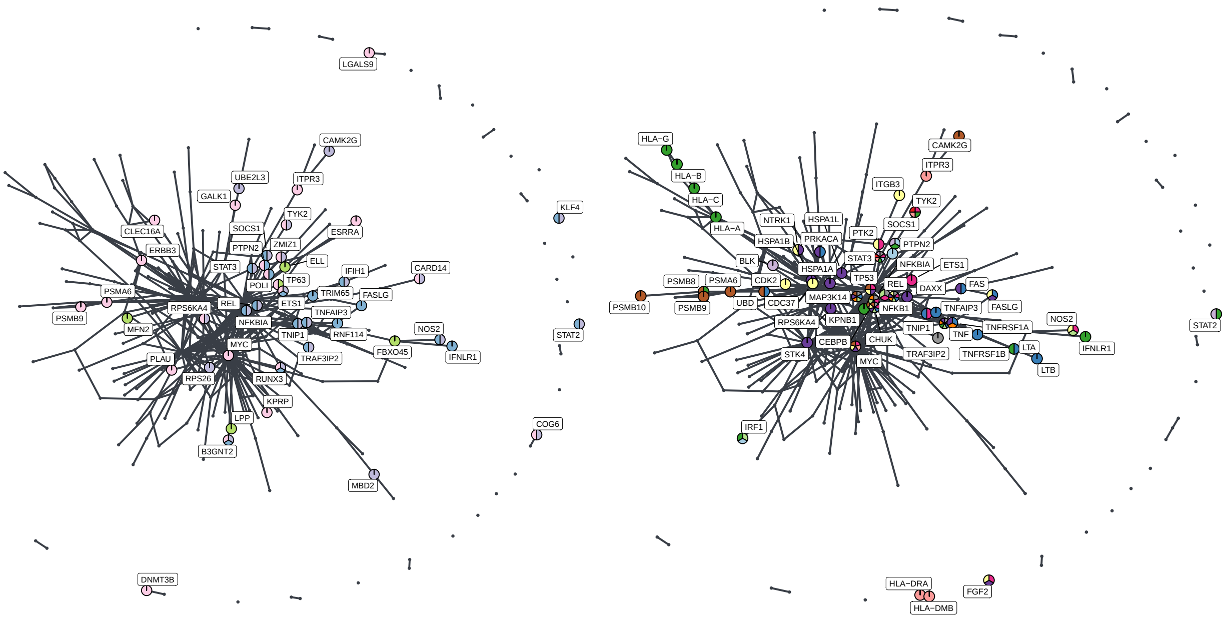


(a) Highlighting otherwise known psoriasis genes.

(b) Highlighting *IL-17*/*IL-23*/*TNF*/*IL-1*/*IFN-γ*/*IFN*-mediated/*IL-22*/*IL-6*/*IL-9*/*NF-κB*/*JAK-STAT*/*MAPK*/*PI3K-AKT*/*Wnt* pathways.

**Fig. S2**: Stable consensus network comprising 258 genes built by choosing those genes selected in at least 7 (a and b) out of 25 solutions. The pie charts show, for each node, which genes are in common with known psoriasis genes (a), and in common with biological pathways found to be associated with psoriasis. In (a) colors represent: Ran et al. (2019)(■), Tsoi et al. (2017) (■), Dand et al. (2025) (■) and Zhang et al. (2025) (■). In (b) colors highlight genes in common with *IL-17* (■), *IL-23* (■), *TNF* (■), *IL-1* (■), *IFN-γ* (■), *IFN*-mediated (■), *IL-22* (■), *IL-6* (■), *NF-κB* (■), *JAK-STAT* (■), *MAPK* (■), *PI3K-AKT* (■) and *WNT* (■) pathways.

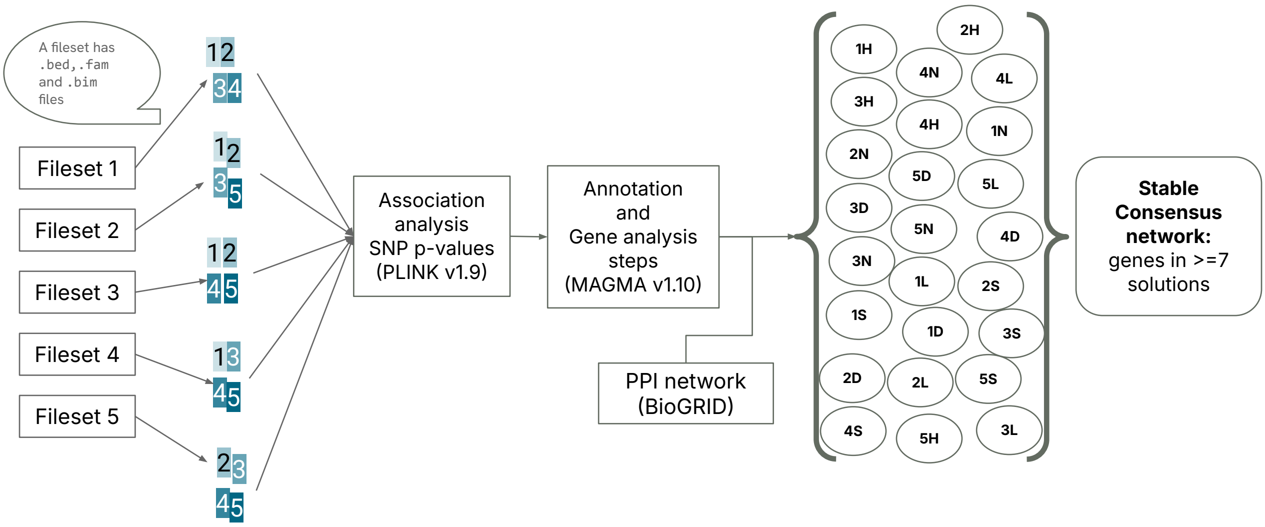


**Fig. S3**: General workflow for constructing the stable consensus network. Each fileset constitutes 80% of the total data samples that have already passed quality control filters. 1H, 2H,.. means the number gene-set solutions produced by Heinz Dittrich et al. (2008), 1N, 2N,.. are the corresponding ones for HotNet2 Leiserson et al. (2015), 1S, 2S,.. are those for SigMod Liu et al. (2017), 1D, 2D, .. correspond to dmGWAS Wang et al. (2015), 1L, 2L, .. correspond to LEAN Gwinner et al. (2017).

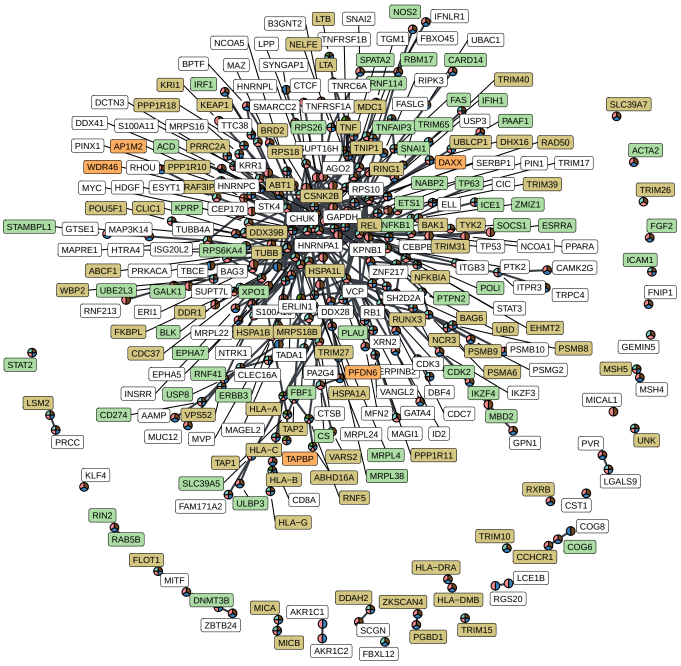


**Fig. S4**: Stable consensus network built by choosing those genes selected in at least 7 out of 25 solutions. There are 258 genes in the network. The pie charts show, for each genes, which method(s) selected it at least one among Heinz (■), HotNet2(■), SigMod(■), dmGWAS(■), and LEAN(■). Labels are colored according to the source its corresponding gene was identified: classical GWAS (cGWAS - ■), MAGMA tool after Benjamini-Hocerg correction (MAGMABH - ■), the intersection of both (MAGMABH and cGWAS - ■) and only using network-methods (□).
